# ALCOHOLIC LIVER BIOMARKERS: DETERMINANTS FOR ADMISSION TO REHABILITATION CENTRES DURING THE COVID 19 PERIOD IN KENYA

**DOI:** 10.1101/2022.10.18.22281212

**Authors:** George Njoroge, Catherine Mwenda, Ezekiel Mecha, Stanley Waithaka

## Abstract

Consumption of alcohol is a practice world over that dates back to 10,000 Before Christ. Evaluation on the damages caused by the alcohol on the human organs such as the liver is paramount. There is no direct evaluation to liver parenchyma before admission rather blood samples are evaluated to show damage or no damage to the liver cells. The outcome of the blood samples denote the health status of the liver cells before admission. Alcohol biomarkers are usually elevated when there is damage to the liver parenchyma. This was a quantitative descriptive cross sectional study. Purpose sampling was used to select two Counties with the highest number of alcohol consumers. Simple Random sampling method was used to select participants for liver biomarkers. Participants were requested to consent for blood donation and confidentiality was maintained. Blood samples collected were separated for serum and cells using centrifuge within one hour after donation. The samples were transported for storage using cool boxes and temperature was maintained between −8 and +8 degrees during transportation. The blood samples were stored at −8 and +8 degrees in the deep freezer. Majority (97%) of the participants had alanine aminotransferase levels of 41 to 80 IU/L. Eighty eight percent of the participants had aspartate aminotransferase elevated to between 35 and 68 IU/L. Gamma Glutamyl aminotransferase was elevated in all of the participants while alanine phosphatase was elevated in 99% of the participants. Most participants had elevated liver biomarkers before admission to rehabilitation centre.

## I. Introduction

Consumption of alcohol over a long duration has negative effects on the functioning of the liver. The liver parenchyma is involved in detoxification of toxins from ethanol. Over time the liver biomarkers are elevated suggestive of liver injury ^1^.

Manthey et al (2019) in a study on 189 countries between 1990 and 2017 found that alcohol drinking had increased from 45% in 1990 to 47% in 2017 with a focus of increasing to 50% by 2030. The interpretation of this statistics was that global reduction of harmful alcohol use was unlikely, the cost of treating the disease was likely to get high. Families and Countries burden for treatment was likely to increase with mortality highly expected to increase. Thus modalities for early detection and early treatment of the disease are important ^2^.

Majority of persons presenting with elevated liver enzymes are asymptomatic ignoring the need for assessment and management of liver injury ^3^. The gold standard in diagnosis of alcoholic liver injury (ALI) focuses on clinical manifestations and laboratory studies ^4^. As such laboratory testing for elevation of the liver biomarkers is needed for definitive diagnosis before admission. The importance of this being to link or exclude the Alcoholic liver injury, with other liver diseases for definitive management ^1,5^.

The diagnosis of ALI largely depends on history from the client upon documentation of the quantity, quality and duration of alcohol consumption ^1,6^.

The history on the amount of alcohol previously consumed may or may not be accurate^7^.

It is highly subjective depending on the circumstance available within the clients which may alter the actual data provided. Disease process may be a contributing factor to erroneous data where a sick person may feel bothered trying to recall the trends of alcohol consumption some of which may date back long durations; therefore, the importance of accurate diagnosis during admission for alcoholic liver injury through blood serum samples ^8^.

### Alcohol consumption during the covid 19 pandemic

> A study in US by Pollard et al (2020) reported during the stay-at-home mitigation measures for coronavirus in 2019 and 2020, the sales of alcohol increased by 54%. The study further reported that online sales for alcoholic products were increased by 262% ^9^.

A study in US by Grossman et al (2020) reported that alcohol consumption had increased during the covid-19 pandemic ^10^. Testino (2020) reported of increase in alcohol consumption during the Covid-19 restrictions ^11^.

A study by Sohal et al (2021) reported that there was a drastic raise in alcohol related hepatitis during the covid 19 pandemic. The study further reported that with the surge in the number of hepatitis patients and the covid 19 patients’ demand for admissions there was need for extra measures to care for alcoholic liver injury patients ^12^.

### Blood serum samples

Laboratory tests to assess liver function is referred to as liver function test, it a clinical practice that involve testing serum samples for bilirubin, alanine aminotransferase, aspartate aminotransferase, alkaline phosphatase, gamma-glutamyltransferase, prothrombin time and albumin. Some tests are more specific to alcohol than others. There is no single liver biomarker that has the capacity to diagnose early alcoholic liver injury or an advanced alcohol liver disease. Therefore the biomarkers complement each other to highly diagnose mal function of the liver ^13^.

Liver enzymatic action is classified as mild when the parameters are less than 5 times Upper Normal Limit (UNL), moderate if the parameters are 5 to 10 times higher on the UNL while severe is classified as more than 10 times UNL. The classification is based on consensus of the UNL and there are no international definitions for these points ^3^.

> A study in US reported on the normal ranges for transaminase values for the biomarkers of liver function as, AST 0 to 35 IU/L, ALT as 0 to 45 IU/L, GGT as 0 to 30 IU/L, Bilirubin as 2 to 17 micromoles/L, Albumin as 40 to 60 g/L and Prothrombin time as 10.9 to 12.5 seconds. Shreevastva, Pandeya and Mishra (2017) reported that AST or ALT when elevated rarely exceed 300 IU/L in ALI ^14,15^.
>
> A study in Kenya by Waithaka et al (2009) showed that ALT range was between 0-39 IU/L, AST was between 6-40 IU/L, ALP was between 13-201, while ALB was between 29-52 IU/L. the study further reported that the difference in range for Kenyan samples were likely due to geographical location of the country, equipments and methods of analysis used and sample sizes for the study ^16^.

Rutayisire (2017) had similar findings where the researcher mentioned that the difference in variance of liver function tests can be influenced by Geographical region and dietary intake ^17^. Eyawo et al (2020) reported of biasness in self reporting from persons of alcohol use ^18^.

### Influence to the blood samples

> Some medications are known to elevate the ALT and AST, Among them are ceftriaxone, cotrimoxazole, phenytoin, carbamazepine and allopurinol. Others include isoniazid, rifampicin, pyrazinamide, dapsone and ibuprofen. Therefore the avoidance of these medication during sample collection is paramount. Periods of increased exercises also raise the levels of transaminase and should be avoided before testing ^19^.

### Statement of the problem

Admission of clients to rehabilitation centres in Kenya is based on history for alcohol consumption. A narration provided by relatives or subjective data from the client on alcohol consumption and chief complain form bases of data for admission to rehabilitation centre. The data is not comprehensive and there is likelihood of biasness. Clinicians use this data to categorize clients for admission or referral based. A more objective method to evaluate on the effects of alcoholic liver injury through liver biomarkers during admission is less involved to eliminate the biasness.

## II. Methodology

The study design was a non-experimental descriptive study design. Sampling Method Due to logistic and resource limitation, purposive sampling method was used to select two counties within Kenya which have high alcohol consumption (NACADA, 2012). The Counties selected were Muranga and Uasin Gishu. Clients within the two counties were stratified into various medical facilities. The medical facilities were then clustered and sampling done. Those medical facilities that was selected were involved in the study. Alcohol consumer clients form a special population and as such all clients in the selected cluster groups of medical facilities were given equal opportunity.

Sample size determination was done using Cochran equation but since the sample was less than 10,000 Yamane formula was used to determine the actual sample which was calculated at 108.

Selection criteria; inclusion criteria involved voluntary consent from clients who accepted voluntary admission to rehabilitation centres while the exclusion criteria, was voluntary withdrawal even after accepting the initial inclusion.

Data Collection Tool. Laboratory investigations forms for Liver Function Tests (LFT) were used. The tests performed included all the tests prescribed in the LFT with a focus to, AST, ALT, GGT, ALP, Direct bilirubin, indirect bilirubin and proteins

Data collection method; Samples of blood were drawn from clients equivalent to 3cc. The samples were drawn by laboratory technologist at the laboratories within the selected medical facilities. Samples were then stored in cool boxes with ice packs. The samples were later transported to Mount Kenya University Innovation and Research Laboratories. They were run through a centrifuge to separate the serum from the erythrocytes. From the serum 0.5mls was drawn and stored in a fridge with temperature readings between −8 and +8 degrees centigrade. The collected blood was put in a vacutainer tube which was labelled using identification code for each participant.

Data Analysis Techniques and Procedures; thawing of the blood samples was done on the day of analysis. Samples were analyzed at the Thika Level 5 hospital laboratory department.

Ethical considerations; The researcher sort for approval from the following institutions; Institutional Ethical Review Committee of Mount Kenya University (approval number 865) and the National Commission for science Technology and innovation (NACOSTI) Kenya (NACOSTI/P/21/10477) which were granted in may 2021.

The study used regression analysis to show relationship between the different parameters for alcohol biomarkers. A test was conducted to establish positive or negative relationship. Analysis of Variance (ANOVA) was used to test for significance

## III. Findings

Data collection was carried out between the months of June and November 2021. Ninety three respondents were involved in the study, this represented 86.11% of the calculated sample size of 108. In our study 96% of the participants were male while 4 percent were female. Regarding age, our study had participants less than 27 years represented by 18% while above the age of 58 years they were represented by 5%.

### Drinking pattern during the Covid-19 restrictions

Regarding alcohol drinking pattern during the Covid-19 period, participants who reported to agree that the covid-19 restrictions affected their alcohol drinking pattern were represented by 44%, participants who reported to disagree that covid-19 restrictions affected their alcohol drinking pattern were represented by 46% while those who reported that covid-19 restrictions partially affected their alcohol drinking pattern were represented by 9.7%.

**Figure 1:**
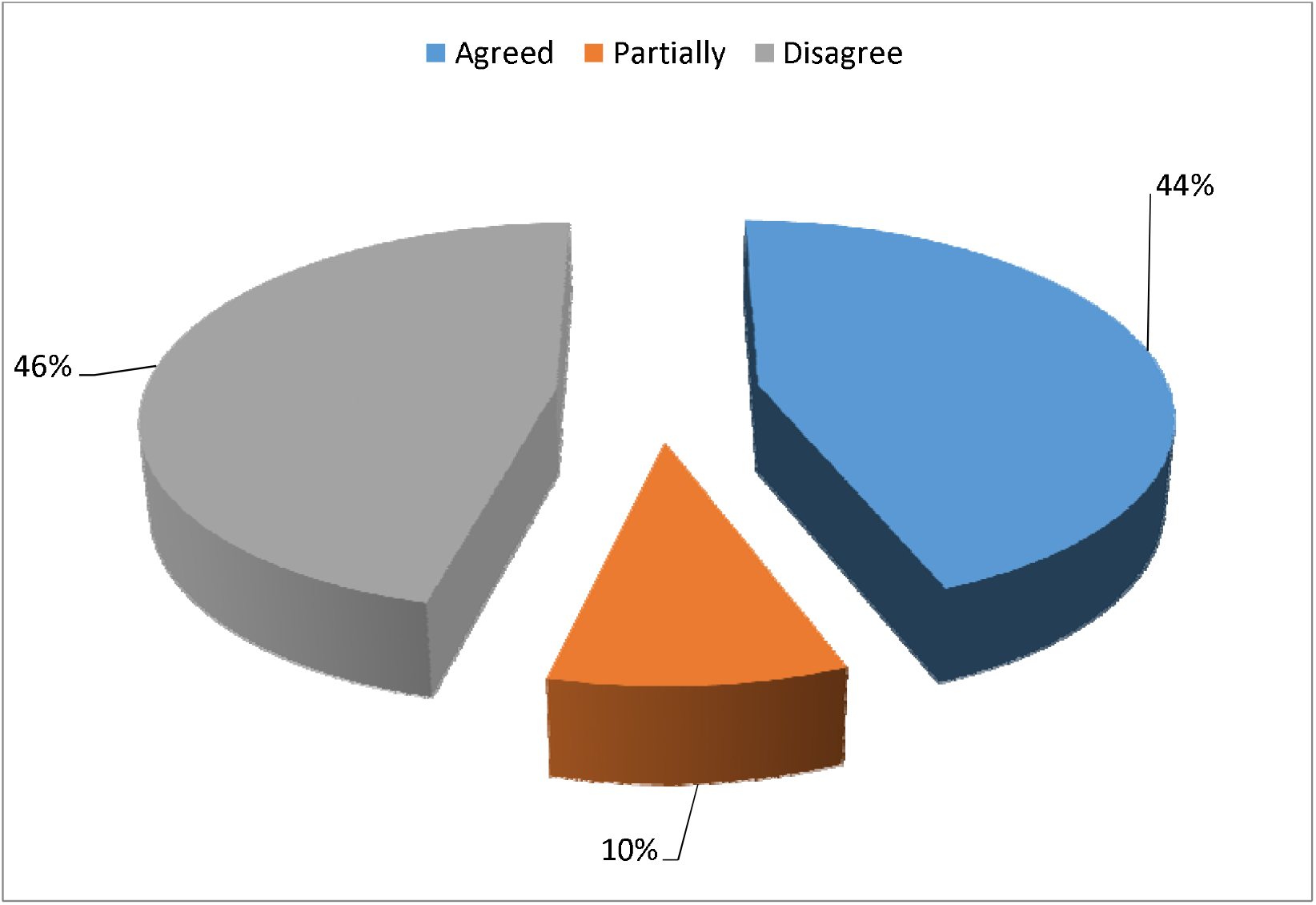
Drinking pattern during the Covid-19 restrictions.

### Descriptive statistics on the levels of alcoholic liver biomarkers among adults of alcohol consumption

**Table 1:**
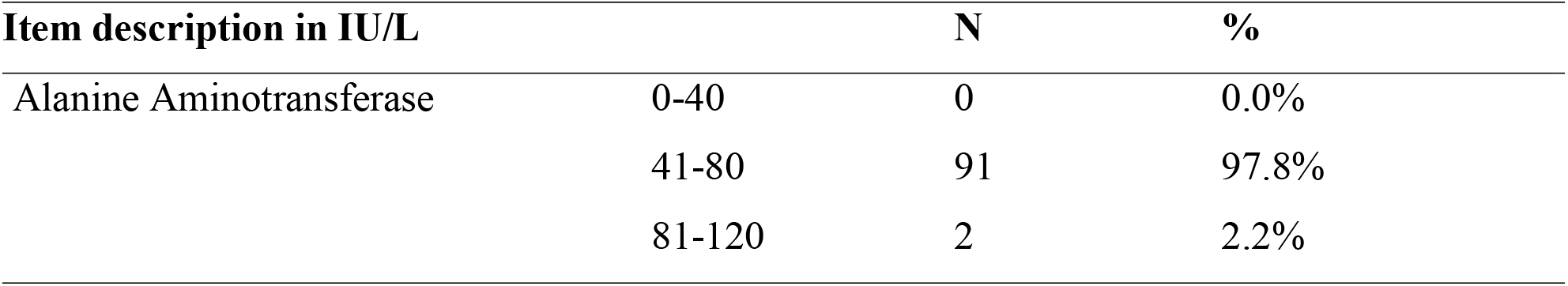
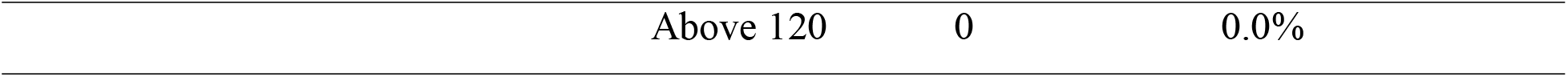
Descriptive Statistics for Aspartate aminotransferase. Alanine aminotransferase diagnostic results

| Item description in IU/L |  | N | % |
| --- | --- | --- | --- |
| Alanine Aminotransferase | 0-40 | 0 | 0.0% |
|  | 41-80 | 91 | 97.8% |
|  | 81-120 | 2 | 2.2% |

|  |  |  |
| --- | --- | --- |
| Above 120 | 0 | 0.0% |

In alanine aminotransferase most of the respondents were found to be between 41 and 80 IU/L represented by 97.8%, followed by those who indicated between 81 and 120 IU/L represented by 2.2% and none (0%) of the respondents were between 0 and 40 IU/L and above 120 IU/L.

**Table 4:**
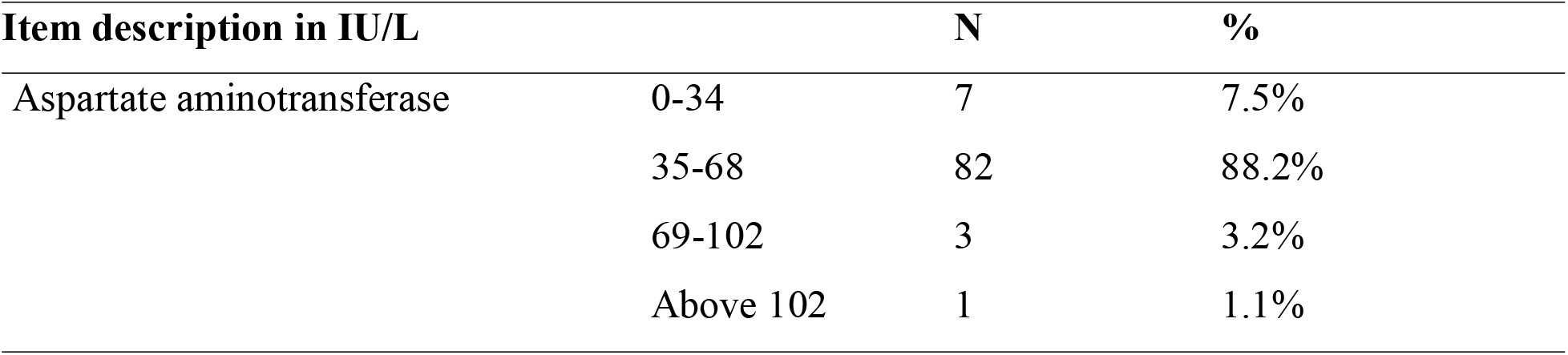
Descriptive statistics for alanine aminotransferase. Aspartate aminotransferase diagnostic results

The research found out that most of the participants had aspartate aminotransferase levels between 35 and 68 IU/L represented by 88.2%, participants with 0 and 34 IU/L were represented by 7.5% while few participants had aspartate levels above 68 IU/L.

**Table 5:** descriptive statistics for Gamma Glutamyl aminotransferase. Gamma glutamyl aminotransferase diagnostic results

| Item description in IU/L |  | N | % |
| --- | --- | --- | --- |
| Gamma Glutamyl aminotransferase | 0-50 | 0 | 0.0% |
|  | 51-100 | 67 | 72.0% |
|  | 101-150 | 26 | 28.0% |
|  | Above 150 | 0 | 0.0% |

In Gamma Glutamyl aminotransferase the research found that more than half of the respondents had levels of between 51 and 100 IU/L represented by 72.0%. Participants with levels between 101 and 150 IU/L were represented by 28.0% while none of the participants had the levels between 0-50 IU/L and above 150 IU/L respectively.

**Table 6:** descriptive statistics for alanine phosphatase. Alanine phosphatase diagnostic results

| Item description in IU/L |  | N | % |
| --- | --- | --- | --- |
| Alanine Phosphatase | 0-150 | 1 | 1.1% |
|  | 151-300 | 86 | 92.5% |
|  | 301-600 | 6 | 6.5% |
|  | above 600 | 0 | 0.0% |

When tested for alanine aminotransferase, almost all the participants had levels between 151 and 300 IU/L represented by 92.5%. Few (6) participants had levels between 301 and 600 IU/L represented 6.5%. Only one (1.1%) had the levels between 0 and 150 while none of the participants had levels above 600 IU/L.

### Regression summary

The model summary was fitted for the various levels of alcoholic liver biomarkers among adults of alcohol consumption it was found out that the value of R^2^ on the relationship between the studied parameters of measuring various level of alcohol consumption was 0.896 showing a goodness of fit of the model. This implied that holding the other factors that are affecting the alcohol consumption constant Alanine Phosphatase, Aspartate aminotransferase, Gamma Glutamyl aminotransferase and Alanine Aminotransferase were main factors that would contribute to liver injury by 89.6%.

**Table 2:** Model summary.

Model Summary
| Model | R | R Square | Adjusted R Square | Std. Error of the Estimate |
| --- | --- | --- | --- | --- |
| 1 | .946<sup>a</sup> | .896 | .896 | .419 |

a. Predictors: (Constant), Alanine Phosphatase, Aspartate aminotransferase, Gamma Glutamyl aminotransferase, Alanine Aminotransferase

**Table 3:** ANOVA Fitting model.

| ANOVA<sup>a</sup> |  |  |  |  |  |  |
| --- | --- | --- | --- | --- | --- | --- |
| Model |  | Sum of Squares | df | Mean Square | F | Sig. |
| 1 | Regression | 3.042 | 4 | .760 | 2.303 | .000<sup>b</sup> |
|  | Residual | .000 | 88 | .033 |  |  |
|  | Total | 3.042 | 92 |  |  |  |

From the four factor which are Alanine Phosphatase, Aspartate aminotransferase, Gamma Glutamyl aminotransferase and Alanine Aminotransferase used to determine the levels of alcoholic liver biomarkers among adults of alcohol consumption it indicated an f-value of 2.303 which was less than the f table value at 4 degree of freedom which was 6.39 hence there was a statistical significance between biomarkers and alcohol consumption. A significance of 0.000 was less than p-value of 0.05 hence showing goodness of fit model.

### Regression analysis

**Table 4:** Regression analysis.

|  |  | Unstandardized Coefficients |  |  |
| --- | --- | --- | --- | --- |
| Model |  | B | Std. Error | Sig. |
| 1 | (Constant) | 4.268 | .000 | 1.000 |
|  | Alanine Aminotransferase | .201 | .000 | .000 |
|  | Aspartate aminotransferase | .536 | .000 | .000 |
|  | Gamma Glutamyl aminotransferase | .620 | .000 | .000 |
|  | Alanine Phosphatase | .372 | .000 | .000 |

The table above shows the regression model on the biomarkers and their corresponding coefficient of each parameter. It was found out that Gamma Glutamyl aminotransferase had the highest influence compare to other factors of 0.620, followed closely by Aspartate aminotransferase with 0.536 correlation rate, Alanine Phosphatase had a correlation of 0.372 and finally Alanine Aminotransferase had a correlation 0.201. We can observe that none of the biomarkers measures had a negative correlation hence positive correlation of biomarkers towards determination of the level of alcoholic liver biomarkers among adults of alcohol consumption.

### Inferential statistics for liver biomarkers

**Table 4:** Paired T-test for Total Bilirubin and Direct Bilirubin

| Paired Samples Correlations |  |  |  |  |
| --- | --- | --- | --- | --- |
|  |  | N | Correlation | Sig. |
| Pair 1 | TBIL & DBIL | 93 | .712 | .000 |

There was a strong correlation between the total bilirubin and the direct bilirubin since there was a high correlation of 71.2% hence can be used to determine the levels of alcoholic liver biomarkers among adults of alcohol consumption. A significance of 0.000 was less than p-value of 0.05 hence showing goodness of fit model.

**Table 5:**
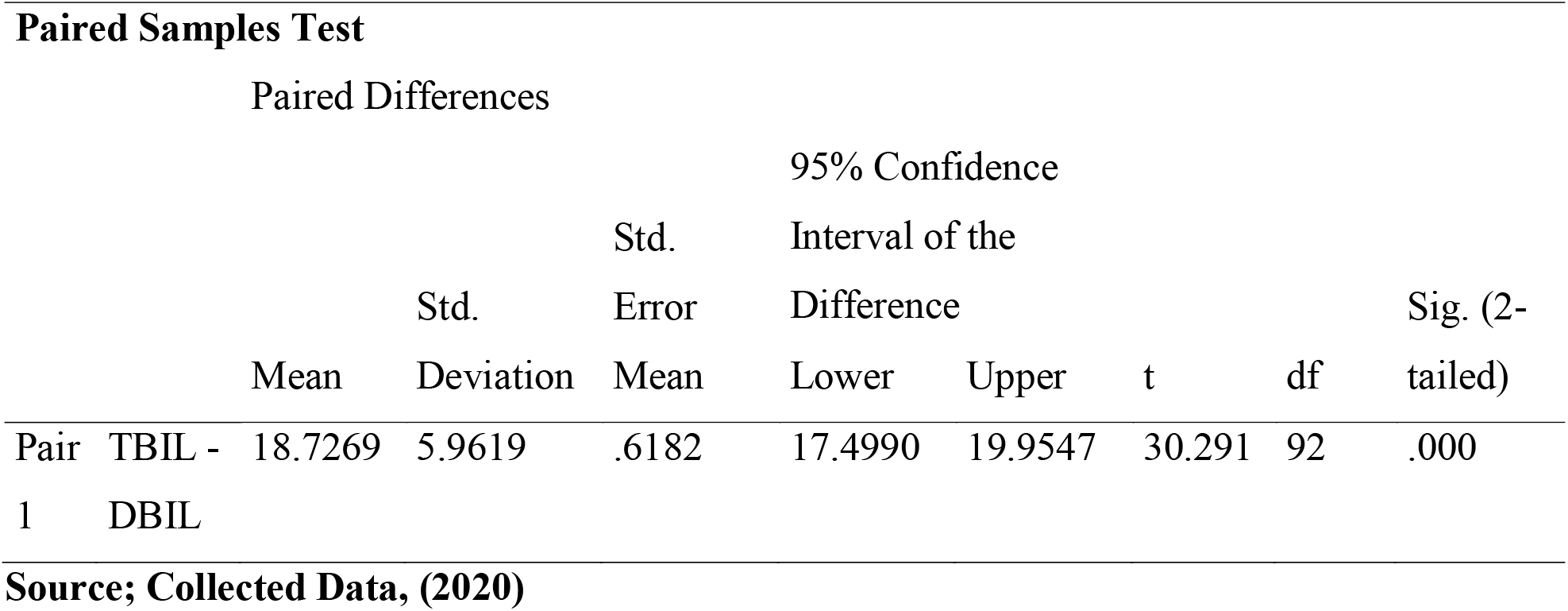
Paired T-test for Total Bilirubin and Direct Bilirubin

A T-value of 30 was obtained from the correlation between the total bilirubin and the direct bilirubin this indicated a strong correlation when compared with the t-table value at 92 degree of freedom which is 1.965 at 95% level of significance.

### Total protein and ALB

Paired sample correlation was carried out and the following results were obtained.

**Table 6:** Paired T-test for Total protein and ALB

Paired Samples Correlations
|  |  | N | Correlation | Sig. |
| --- | --- | --- | --- | --- |
| Pair 1 | TP & ALB | 93 | .56 | .000 |

There was half of correlation between the total protein and the ALB with a correlation of 56.0% hence indicating there was a relationship between the total protein and the ALB. A significance of 0.000 was obtained which is less than 0.05 this indicating that goodness of fit in the model.

Paired Samples Test
|  |  | Paired Differences |  |  | 95% Confidence |  |  |  |  |
| --- | --- | --- | --- | --- | --- | --- | --- | --- | --- |
|  |  |  |  | Std. | Interval of the |  |  |  |  |
|  |  | Mean | Std. Deviation | Error Mean | Difference |  | T | df | Sig. (2-tailed) |
| Pair 1 | TP - ALB | 29.077 | 9.787 | 1.015 | 27.062 | 31.093 | 28.652 | 92 | .000 |

Paired T-test between the total protein and the ALB obtained a t value of 28.652 which is greater than T-table value which is 1.965 at 95% level of significance at 92 degree of freedom. This indicated that there was an association between the total protein and the ALB in estimating the levels of alcoholic liver biomarkers among adults of alcohol consumption.

### Correlation of the Total protein, Alanine Aminotransferase, Aspartate aminotransferase Gamma Glutamyl aminotransferase and Alkaline phosphatase

The table below indicates the results of the correlation between where Total protein was compared with the Alanine Aminotransferase, Aspartate aminotransferase Gamma Glutamyl aminotransferase and Alkaline phosphatase.

**Table 7:** Correlation of the Total protein, Alanine Aminotransferase, Aspartate aminotransferase Gamma Glutamyl aminotransferase and Alkaline phosphatase

| Paired Samples Correlations |  |  |  |  |
| --- | --- | --- | --- | --- |
|  |  | N | Correlation | Sig. |
| Pair 1 | TP & ALT | 93 | -.062 | .000 |
| Pair 2 | TP & AST | 93 | -.050 | .000 |
| Pair 3 | TP & ALP | 93 | -.043 | .000 |
| Pair 4 | TP & GGT | 93 | .071 | .000 |

The paired value correlation between the total protein and the Alanine Aminotransferase was − 0.062 indicating a negative correlation relationship between the Total protein and ALT. The relationship between the Total protein and the Aspartate aminotransferase had a negative correlation of −0.050 hence contributing levels of alcoholic liver biomarkers among adults of alcohol consumption. Correlation of −0.043 was drawn between the relationship between the total protein and Alkaline phosphatase similarly indicating a correlation hence negative impacts towards the influence towards contributing levels of alcoholic liver biomarkers among adults of alcohol consumption. The correlation between the total protein and the Gamma Glutamyl aminotransferase was very high with 0.071 hence indicating a statistical relationship between them hence influence towards contributing levels of alcoholic liver biomarkers among adults of alcohol consumption. A significance of 0.000 indicated the goodness in fit of the model in paired t-test.

### Paired T-test for the Total protein, Alanine Aminotransferase, Aspartate aminotransferase Gamma Glutamyl aminotransferase and Alkaline phosphatase

The table below indicates the results of the paired T-test between where Total protein was compared with the Alanine Aminotransferase, Aspartate aminotransferase Gamma Glutamyl aminotransferase and Alkaline phosphatase.

**Table 8:**
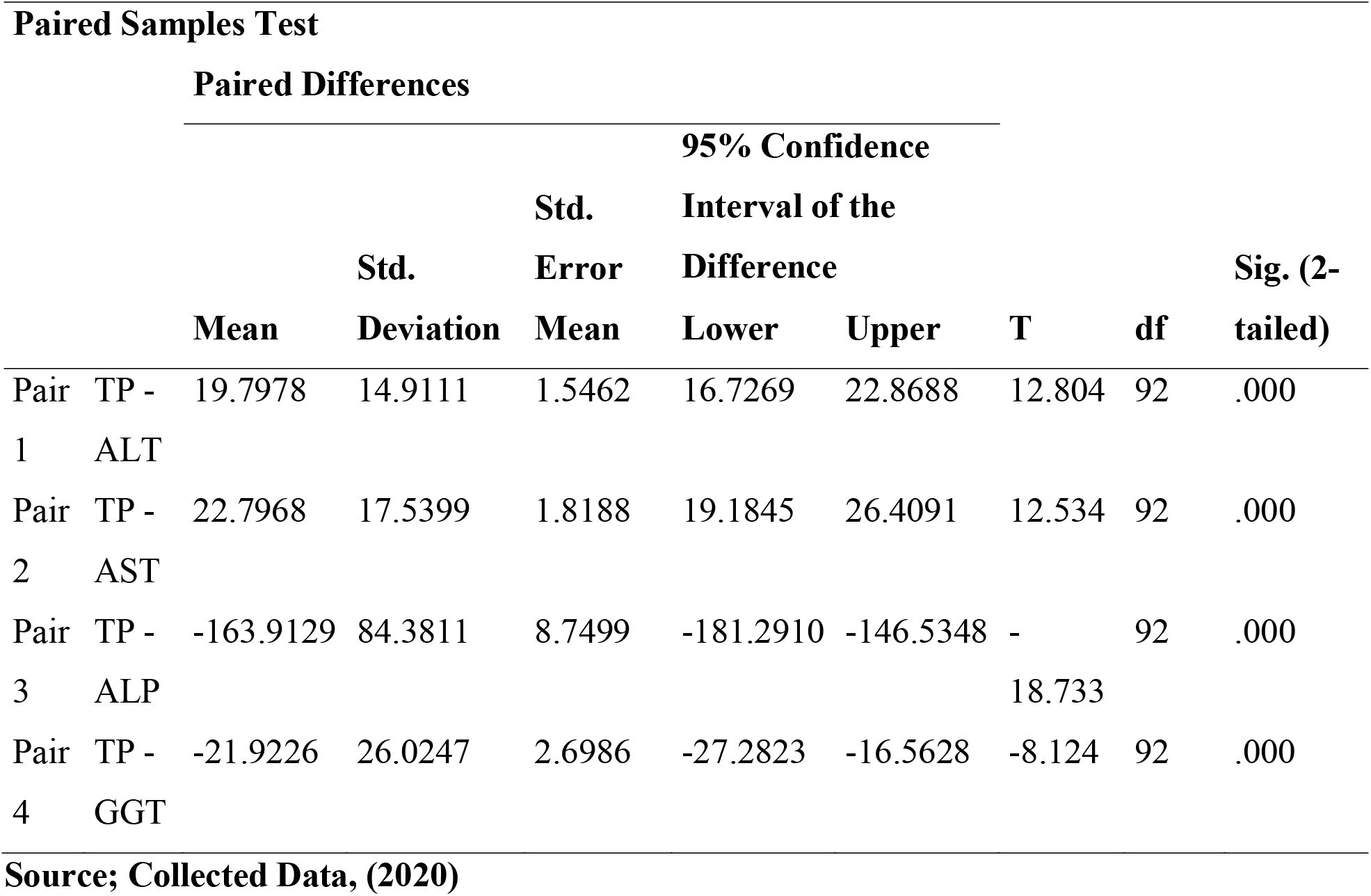
Paired T-test for the Total protein, Alanine Aminotransferase, Aspartate aminotransferase Gamma Glutamyl aminotransferase and Alkaline phosphatase

A t-value of 12.804 was obtained between the pair of the total protein and the Alanine Aminotransferase, this indicated a strong correlation between them since the table value is 1.66 at 92 degree of freedom at 95% confidence interval. The paired value between the Total protein and the Aspartate aminotransferase was 12.534 which indicating a relationship between the two variables hence influence towards contributing levels of alcoholic liver biomarkers among adults of alcohol consumption at 95% confidence interval with a t-value of 1.66.

The relationship between the total protein and Alkaline phosphatase was 18.733 this indicated a relationship between the total protein and Alkaline phosphatase towards the influence towards contributing levels of alcoholic liver biomarkers among adults of alcohol consumption at 95% confidence interval. A t-value of −8.124 was generated between the total protein and the Gamma Glutamyl aminotransferase this is less than the t-table value at 92 degree of freedom at 95% confidence

## IV. Discussion

This study was done during the covid-19 restrictions and as such there was need to evaluate the admission criteria in rehabilitation centres. Majority of the participants were male with few female, suggestive that many male were ready to stop alcohol consumption and journey through abstinence for recovery.

Few female may be suggestive of fewer number of female involved in admission to rehabilitation centres due to alcohol consumption than male. The findings were in consistent with those from Uganda with a near geographical region to Kenya and those from USA where few female than men were recruited to rehabilitation centres ^20, 21^.

Regarding age, our study had diferrent findings from Coetzee, et al (2022) who reported that majority (45%) of the participants for admission due to alcoholic liver injury were between the age of 18to 29 years ^22^. In other studies admission due to ALI had a median age of 51 years ^23^.

### Drinking pattern during the Covid-19 restrictions

The findings on this study showed that nearly half of the participants disagreed that covid-19 restrictions affected their alcohol drinking pattern while nearly half of participant agreed that covid-19 restrictions affected their alcohol drinking pattern.

#### Liver biomarkers

Our study found that majority of the participants had elevated liver biomarkers suggestive of impending or existing liver injury prior to admission. The results were supported by the participants need to sort medical attention at the rehabilitation centres during the covid 19 period. Pott-Junior et al (2019) had similar findings where the liver biomarkers were elevated during admissions with follow-up laboratory evaluation during the hospital stay^24^.

Our study showed GGT as the main biomarker of influence among the tested alcoholic biomarkers. These finding were consistent with those from a study in India by Ghosh et al (2019) which reported the importance of GGT as a dependable biomarker during admission ^25^.

Contrary opinion was sited by Moringa et al (2022) who reported of other parameters in LFTs such as Carbohydrate-deficient transferrin which were more sensitive than GGT ^26^.

Others were in the following order AST, AP and then ALT. There was a correlation in elevation between AST and ALT where AST was elevated similarly ALT was elevated. Sutoh et al (2020) and Wu et al (2020) reported of an association between aspartate aminotransferase and alanine aminotransferase with both parameters elevated upon rise of the other ^27,28^.

The findings showed the need for every client to be evaluated for liver injury during admission to rehabilitation centres with suggestions for further evaluation. A study in India by Anttila et al (2005) had similar findings where it was suggested of subsequent liver biomarkers evaluation until discharge or referral^29^. The findings of the study suggested GGT, AST and ALT as the main biomarkers for severity of liver injury. Different studies have also reported of these parameters as important predictors of mortality due to chronic alcohol consumption ^30, 31^.Other researchers had contrary opinions from our study citing no importance in liver function tests during admission and emphasized need for behavioral approach in treating alcoholism than systemic approach ^32,33,34^

## Conclusion

This study emphases the importance of diagnostic evaluation during admission to rehabilitation centres as a norm to good practice of medical care. Clients are better managed with baseline information about the severity of alcoholic liver injury. In incidences of improvement or deterioration the admission data is of paramount importance to guide the clinicians for referral or otherwise.

### Gender

Male were majority of the participants in the study with few female. There were predictions to the availability of more men than women ready for admission, since more men were involved in alcohol consumption than female. The stigma of women and alcohol consumption had an influence on women presenting themselves on the rehabilitation centres. More men were probably on the focus from the community to cut down on alcohol than female.

### Alanine aminotransferase

This biomarker of the liver was elevated in almost all the participants. Few (2) had high levels which may have showed advanced levels of liver damage. Otherwise all the participants had elevated ALT showing there was alcohol liver injury in almost all the participants.

### Aspartate Aminotransferase

It is probably the most dependable biomarker of all. Its elevation in alcohol consumers is dependable to alcoholic liver injury. In our study the biomarker was elevated with few (7) participants reporting normal results.

### Gamma Glutamyl aminotransferase

This is another dependable biomarker of the liver. It was elevated in all the participants. Again when combined with other liver biomarkers it shows alcoholic liver injury. Our participants were likely cases of morbidity due to alcoholic liver injury.

### Alkaline phosphatase

This biomarker was elevated on most participants with an exception of only one participant. Majority had elevated levels showing that it is true their alcohol consumption had effect on the liver function.

### Recommendations

All clients due for admission to rehabilitation centres due to alcoholic liver injury should undergo baseline evaluation for liver biomarkers. The findings should guide the subsequent care for the stay or referral of the client.

### Gender

More studies should be done to find out whether more men were involved in alcohol consumption than female, whether there is stigma of the female gender when they present themselves at the rehabilitation centres or any other study that can elicit more information on the skewness of more male than female at the rehabilitation centres.

### Alanine aminotransferase

The biomarker is a dependable test for admission due to alcoholic liver injury that is more reliable when combined with other biomarkers. The study recommended that this biomarker be combined with other biomarkers to provide high level diagnosis for alcoholic liver injury during admission to rehabilitation centres.

### Alanine aminotransferase

It is among the elevated biomarkers of alcoholism and should form part of the diagnostic evaluation criteria during admission for alcoholic liver injury.

### Gamma Glutamyl aminotransferase

Being a dependable biomarker of alcoholic liver injury, it should be combined with other biomarkers to offer more diagnosis evaluation for alcoholic liver injury. The biomarker should form a part of the admission criteria for alcoholic liver injury.

### Alkaline phosphatase

Alkaline phosphatase is also dependable as it was elevated in most participants. It should form part of the liver function tests for evaluation of alcoholic liver injury before admission.

## Data Availability

All data produced in the present study are available upon reasonable request to the authors

## Declarations

### List of abbreviations

ADH: Alcohol Dehydrogenase
ALD: Alcoholic liver Disease
ALDH: Aldehyde Dehydrogenase
ALI: Alcoholic Liver Injury
ALT: Alanine Aminotransferase
ALP: Alkaline Phosphatase
AST: Aspartate Aminotransferase
AUD: Alcohol Use Disorder
BC: Before Christ
CDT: Carbohydrate Deficient Transferin
EASL: European Association for the Study of Liver
GGT: Gamma-Glutamyl Transferase
IU/L: International Units per Litre
IU: International Units
LFT: Liver Function Tests
MKU: Mount Kenya University
NACADA: National Alcohol and Drug Abuse
NACOSTI: National Commission for Science, Technology and Innovation
NALD: Non Alcoholic Liver Disease
PEth: Phosphatydilethanol
SPSS: Statistical Package for Social Science
UK: United Kingdom
US: United States
USA: United State of America
WHO: World Health Organization

## Ethics approval and consent to participate

The researcher sort for approval from the following institutions; Institutional Ethical Review Committee of Mount Kenya University (approval number 865) and the National Commission for science Technology and innovation (NACOSTI) (NACOSTI/P/21/10477).

## Consent for publication

Not applicable

## Availability of data and materials

Data and materials for this manuscript will be available upon request from the author

## Authors’ contributions

Njoroge George Kimani, was the main author of the study

Catherine Syombua Mwenda, was the co-author of the study

Ezekiel Mecha, was the co-author of the study

Stanley Waithaka, was the co-author of the study

## Acknowledgements

This study acknowledges the contribution of rehabilitation centres within Muranga and Uasin Gishu counties in Kenya. We also acknowledge the support from Mount Kenya University Research and Innovation centre for storage facilities and the Thika level five hospital for laboratory work-ups.

## Authors’ information

Njoroge George Kimani

Mount Kenya University

School of Nursing

Catherine Syombua Mwenda

School of Nursing Sciences and Public Health

South Eastern Kenya University

Ezekiel Mecha

School of Biochemistry

University of Nairobi

Stanley Waithaka

School of Medicine

Mount Kenya University

